# Hearing loss at 6-monthly assessments from age 12 to 36 months: secondary outcomes from randomised controlled trials of novel pneumococcal conjugate vaccine schedules

**DOI:** 10.1101/2024.03.13.24304198

**Authors:** A.J. Leach, N. Wilson, B. Arrowsmith, J. Beissbarth, E.K. Mulholland, M. Santosham, P.J. Torzillo, P. McIntyre, H. Smith-Vaughan, S.A. Skull, V.M. Oguoma, M.D. Chatfield, D. Lehmann, C. G. Brennan-Jones, M. J. Binks, P.V. Licciardi, R. Andrews, T. Snelling, V. Krause, J. Carapetis, A.B. Chang, P.S. Morris

## Abstract

**Introduction:** In remote communities, Australian First Nations children with hearing loss are disproportionately at risk of poor school readiness and performance, compared to those with normal hearing. Our objective was to compare two pneumococcal conjugate vaccine (PCV) formulations and mixed schedules (the PREVIX trials) designed to broaden protection and reduce conductive hearing loss to age 36 months.

**Methods:** In two sequential parallel, open-label, randomised controlled trials, eligible infants were first allocated 1:1:1 at age 28-38 days to standard or mixed PCV primary schedules, then at age 12 months to a booster dose (1:1) of PCV13 (13-valent pneumococcal conjugate vaccine, +P) or PHiD-CV10 (10-valent pneumococcal *Haemophilus influenzae* protein D conjugate vaccine, +S). Here we report secondary hearing outcomes in the +P and +S groups at 6-monthly scheduled assessments from age 12 to 36 months.

**Findings:** From March 2013 to September 2018, 461 hearing assessments were performed. Prevalence of mild-moderate hearing loss declined in both groups from ∼75% at age 12 months to ∼53% at 36 months. At primary endpoint age 18 months, prevalence of moderate (disabling) hearing loss was 21% and 41% in the +P and +S groups, respectively (difference −19% [95% confidence interval −38, −1], p=0.07) and prevalence of normal hearing was 36% and 16%, respectively (difference 19% [95%CI 2, 37], p=0.05). At subsequent timepoints prevalence of moderate hearing loss remained lower in the +P group at −3% [95% CI −23, 18] at age 24 months, −12% [95%CI −30, 6] at 30 months, and −9% [95%CI −23, 5] at 36 months.

**Interpretation:** This study provides first evidence of the high prevalence and persistence of mild and moderate hearing loss throughout early childhood. A lower prevalence of moderate (disabling) hearing loss in the +P group may have substantial benefits for high-risk children and warrants further investigation.

**Trial registration:** ClinicalTrials.gov NCT01735084 and NCT01174849 https://clinicaltrials.gov/study/NCT01735084 https://clinicaltrials.gov/study/NCT01174849

**Funding:** National Health and Medical Research Council of Australia (GNT605810, GNT1046999, GNT1120353)

## Introduction

### Background and objectives

Australian First Nations children living in remote communities continue to experience social and educational disadvantage[1] which can be attributed in part to preventable hearing loss associated with early onset of persistent otitis media (OM). *Streptococcus pneumoniae* (pneumococcus) and non-typeable *Haemophilus influenzae* (NTHi) are dominant pathogens of OM from soon after birth.[2]

The Northern Territory (NT) childhood vaccination schedule replaced seven-valent pneumococcal conjugate vaccine (PCV7) with ten-valent pneumococcal *Haemophilus influenzae* protein D conjugate vaccine (S, PHiD-CV10) in 2009, then 13-valent PCV (P, PCV13) vaccine in 2011. Our surveillance studies found evidence of less AOM and a lower prevalence of NTHi in ear discharge of PHiD-CV10-vaccinated compared to PCV7-vaccinated First Nations children.[2, 3]

Our objective was to conduct two sequential randomised controlled trials (RCTs) of primary- and booster-head-to-head and combination schedules of PCV13 and PHiD-CV10; PREVIX_COMBO[4, 5] and PREVIX_BOOST,[6] PREVIX being a name representing the two PCV brand names.

We hypothesised that infants receiving PHiD-CV10 compared with those receiving PCV13 as a booster at 12 months of age would have (i) superior immunogenicity at 18 months of age for common serotypes (1, 4, 5, 6B, 7F, 9V, 14, 18C, 19F and 23F), and at ages 18, 24, 30 and 36 months of age, infants receiving PHiD-CV10 compared with those receiving PCV13 as a booster at 12 months of age would have (ii) less nasopharyngeal carriage of NTHi; (iii) more NP carriage of serotypes 3, 6A and 19A; (iv) less NP carriage of common serotypes (1, 4, 5, 6B, 7F, 9V, 14, 18C, 19F and 23F); (v) less NP carriage of vaccine-related or nonvaccine (replacement) serotypes; (vi) less OM; (vii) less respiratory illness; (viii) less hearing loss and (ix) less developmental delay.[7] We have reported co-primary and select secondary outcomes to age 18 months,[4, 6, 8] and otitis media to age 36 months.[9]

This study, PREVIX_VOICES (Vaccines for Otitis In Children Entering School) builds on PREVIX_BOOST through added capacity to employ paediatric audiologists to conduct 6-monthly hearing tests from age 12 to 36 months. Here we report new data on vaccine group comparisons of hearing level and prevalence of hearing loss.

## Methods

Details of PREVIX_COMBO and PREVIX_BOOST (including PREVIX_VOICES) protocols have been published.[5, 7] Brief methods relevant to hearing outcomes from age 12 to 36 months are described below.

### Study design

The PREVIX_COMBO and PREVIX_BOOST trials were primary outcome assessor-blinded, three-arm (1:1:1) and two-arm (1:1) parallel RCTs. PREVIX_VOICES added capacity to obtain the hearing outcomes for all children in the PREVIX_BOOST study, and additional scheduled 6-monthly assessments at ages 24 to 30 months. No changes that affected the primary objectives or outcomes were made after trial commencement.

### Participants

First Nations children were eligible for PREVIX_BOOST if they were 12 months of age, had previously enrolled in the PREVIX_COMBO three-arm RCT, and were living in one of three remote Aboriginal communities in the NT or a single Western Australian community. PREVIX_VOICES evaluated hearing outcomes in all PREVIX-enrolled children living in the three NT communities.

### Randomisation and masking

Sequence generation: After confirming eligibility and obtaining informed consent, research nurses called the 24/7 randomisation service of the National Health and Medical Research Council Clinical Trial Centre to obtain allocation. Stratification by remote community was applied in both trials, and for PREVIX_BOOST, minimisation by PREVIX_COMBO group was applied. This open-label design did not allow for blinding.

### Procedures

PHiD-CV10 contains 10 pneumococcal serotypes 1, 4, 5, 6B, 7F, 9V, 14, 18C, 19F, 23F and incorporates protein D of NTHi as a carrier protein for eight serotypes. PCV13 contains these ten and additional serotypes 3, 6A, and 19A, but does not incorporate protein D.

Audiology assessments were initially made by fly-in/fly-out NT Government Hearing Health Program audiologists. Paediatric audiologists dedicated to assessment of hearing in the PREVIX_VOICES study commenced in March 2017. All assessments were conducted in the child’s community, in sound-treated hearing booths using an audiometer (Interacoustics paediatric audiometer (PA5) or Otometrics MADSEN Itera II). Hearing responses were tested using Visual Reinforcement Observation Audiometry (VROA) in the sound-field or Play Audiometry using headphones. The pure-tone four-frequency average (4fa) hearing level (in decibels, dB HL) was calculated for air conduction thresholds binaural or monaurally using up to four frequencies 0·5, 1·0, 2·0, and 4·0 kHz (i.e. ≥4fa). Categories of hearing response (hereafter referred to as hearing loss) were no or slight hearing loss (≥ 0 to 20 dB), mild (21 to 30 dB), moderate (31 to 60 dB), severe (61 to 90 dB), or profound (>90dB) hearing loss.[10] We also report the proportion with mean hearing level (3fa) greater than 23·3 dB, which is the Hearing Australia threshold for hearing assistance.[11] All child-visits included a general health check, with management according to local guidelines.[12, 13]

### Analysis and statistical methods

Sample sizes of 425 and 270 were estimated for primary outcomes (immunogenicity) of the PREVIX_COMBO and PREVIX_BOOST, respectively.[4, 6] Analyses of secondary outcome data available at each time point are according to allocated group. We did not impute any data. All data were analysed using Stata/IC version 15.1[14] Baseline characteristics are reported for each PREVIX_BOOST group using means and standard deviation for continuous data if assumption of normal distribution was met, otherwise median value and IQR are reported. Categorical data are summarised as percentages. Vaccine group differences in percentages are compared with two-sided Clopper-Pearson 95% Confidence Interval (95%CI, Fisher’s Exact p value) and differences in mean hearing level as two-sample t-test with equal variances.

## Results

The PREVIX_BOOST trial commenced participant recruitment in March 2013. Hearing tests were performed by the NT Government Hearing Services outreach program if possible. Research audiologists commenced in March 2017. Data collection was completed in September 2018.

At baseline age 12 months, 131 children were allocated to +P and 130 to +S; one child allocated to +P received +S and was included in the +P group for analysis. The numbers of children seen and assessed for hearing outcome are given in Fig 1 and Table 1. Of 461 hearing tests, 409 (89%) were VROA and 52 (11%) Play Audiometry; 74% were performed by research audiologists. Due to the late start of research audiologists, only 23% to 35% PREVIX_BOOST children were age-eligible for the 12- and 18-month hearing assessments, respectively, and around half (58%) had hearing tests at age 36 months. For the additional study visits at ages 24 and 30 months, almost all age-eligible children had hearing assessments (Table 2, Fig 1).

**Fig 1.**
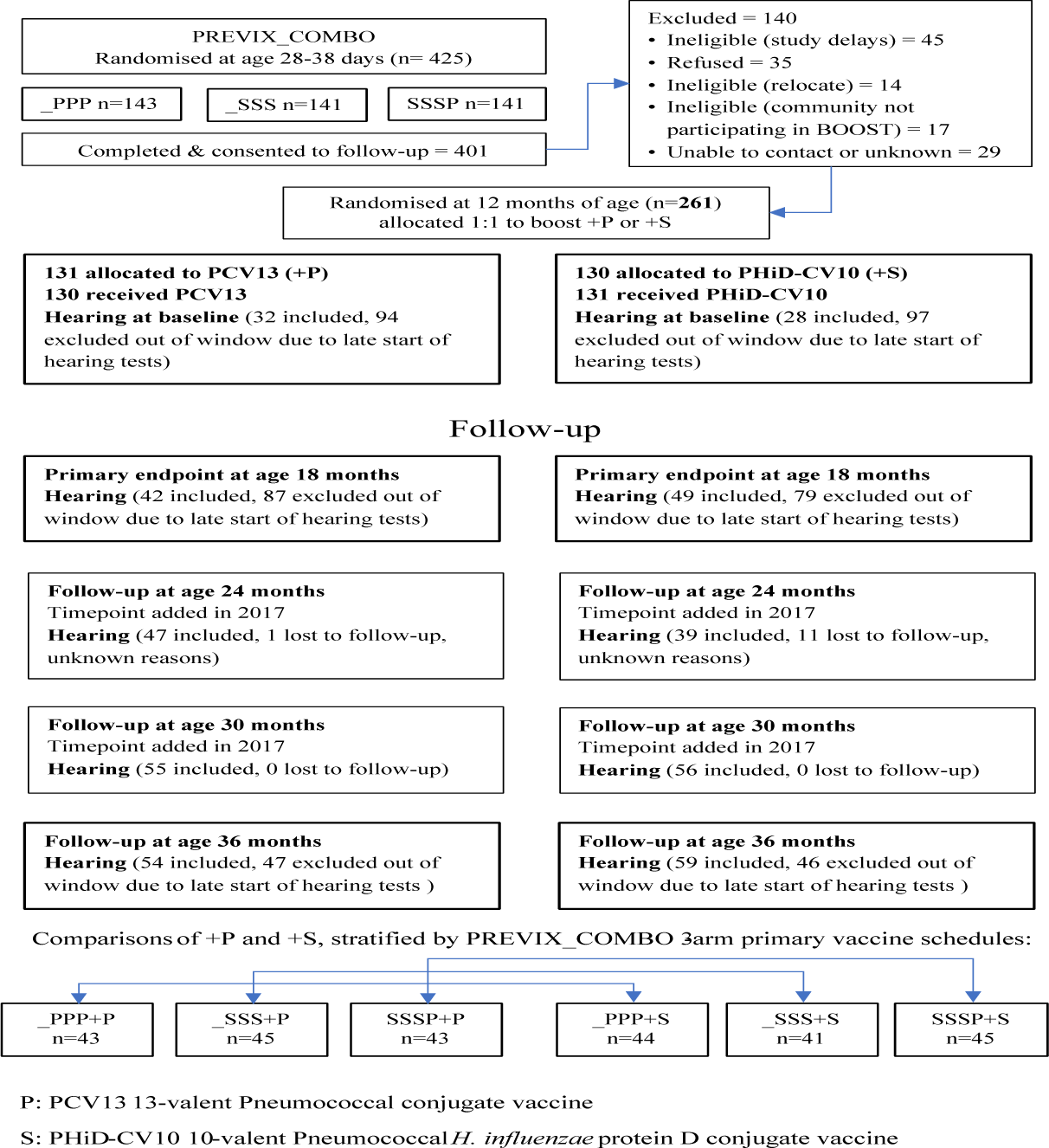
Participant Flow.

**Table 1.** Baseline characteristics and risk factors for the hearing cohort at each study visit age.

| **Vaccine group** |  | **+P** | **+S** | **+P** | **+S** | **+P** | **+S** | **+P** | **+S** | **+P** | **+S** |
| --- | --- | --- | --- | --- | --- | --- | --- | --- | --- | --- | --- |
| **Age at hearing test, months (SD)** |  | **12·7**<br>**(1·1)** | **12·6**<br>**(1·4)** | **18·4**<br>**(1·1)** | **18·5**<br>**(1·4)** | **24·1**<br>**(1·2)** | **24·3**<br>**(1·2)** | **30·2**<br>**(1·5)** | **30·3**<br>**(1·4)** | **36·9**<br>**(1·9)** | **36·9**<br>**(1·8)** |
| **N** |  | **32** | **28** | **42** | **49** | **47** | **39** | **55** | **56** | **54** | **59** |
| Sex | male | 16/32<br>(50%) | 14/28<br>(50%) | 19/42<br>(45%) | 24/49<br>(49%) | 22/47<br>(47%) | 19/39<br>(49%) | 29/55<br>(53%) | 36/56<br>(64%) | 27/54<br>(50%) | 37/59<br>(63%) |
| Gestational age at birth, weeks (SD) |  | 38·2<br>(1·2) | 38·1<br>(2·0) | 38·3<br>(1·2) | 38·0<br>(1·7) | 37·9<br>(1·5) | 38·2<br>(1·7) | 38·1<br>(1·2) | 38·1<br>(1·5) | 38·0<br>(1·5) | 38·2<br>(1·5) |
| Birth weight, Kg (SD) |  | 3·08<br>(0·48) | 3·17<br>(0·49) | 3·06<br>(0·49) | 3·17<br>(0·49) | 3·13<br>(0·56) | 3·13<br>(0·44) | 3·02<br>(0·48) | 3·11<br>(0·46) | 3·14<br>(0·52) | 3·12<br>(0·38) |
| Any otitis media (otoscopy and tympanometry) | Yes | 25/30<br>(83%) | 25/26<br>(96%) | 38/41<br>(93%) | 43/48<br>(90%) | 31/42<br>(74%) | 31/35<br>(89%) | 39/53<br>(76%) | 40/50<br>(80%) | 30/51<br>(59%) | 39/56<br>(70%) |
| Were you breastfeeding? | Yes | 31/32<br>(97%) | 25/27<br>(93%) | 30/35<br>(86%) | 34/41<br>(83%) | 38/46<br>(83%) | 30/39<br>(77%) | 37/55<br>(67%) | 43/55<br>(78%) | 28/54<br>(52%) | 43/59<br>(73%) |
| Community | Wurrumiyanga | 11/32<br>(34%) | 8/28<br>(29%) | 10/42<br>(24%) | 14/49<br>(29%) | 15/47<br>(32%) | 12/39<br>(31%) | 13/55<br>(24%) | 16/56<br>(29%) | 11/54<br>(20%) | 16/59<br>(27%) |
|  | Wadeye | 14/32<br>(44%) | 13/28<br>(46%) | 22/42<br>(52%) | 20/49<br>(41%) | 20/47<br>(43%) | 14/39<br>(36%) | 21/55<br>(38%) | 23/56<br>(41%) | 26/54<br>(48%) | 26/59<br>(44%) |
|  | Maningrida | 7/32<br>(22%) | 7/28<br>(25%) | 10/42<br>(24%) | 15/49<br>(31%) | 12/47<br>(26%) | 13/39<br>(33%) | 21/55<br>(38%) | 17/56<br>(30%) | 17/54<br>(31%) | 17/59<br>(29%) |
| How many children under age 5 years live with you and your baby? | 0 | 14/32<br>(44%) | 11/28<br>(39%) | 18/42<br>(43%) | 20/49<br>(41%) | 25/47<br>(53%) | 20/39<br>(51%) | 28/55<br>(51%) | 23/56<br>(41%) | 20/54<br>(37%) | 28/59<br>(47%) |
|  | 1 | 10/32<br>(31%) | 5/28<br>(18%) | 15/42<br>(36%) | 13/49<br>(27%) | 12/47<br>(26%) | 12/39<br>(31%) | 14/55<br>(25%) | 18/56<br>(32%) | 17/54<br>(31%) | 18/59<br>(31%) |
|  | >1 | 8/32<br>(25%) | 12/28<br>(43%) | 9/42<br>(21%) | 16/49<br>(33%) | 10/47<br>(21%) | 7/39<br>(18%) | 13/55<br>(24%) | 15/56<br>(27%) | 17/54<br>(31%) | 13/59<br>(22%) |
| Have any of your other children had runny ears? | Yes | 3/21<br>(14%) | 3/23<br>(13%) | 7/33<br>(21%) | 10/36<br>(28%) | 5/40<br>(13%) | 10/31<br>(32%) | 9/45<br>(20%) | 13/43<br>(30%) | 12/49<br>(24%) | 8/48<br>(17%) |
| Has your baby had any bad runny nose since the last study visit? | Yes | 12/32<br>(38%) | 12/28<br>(43%) | 19/42<br>(45%) | 29/48<br>(60%) | 19/47<br>(40%) | 17/39<br>(44%) | 27/55<br>(49%) | 21/56<br>(38%) | 23/53<br>(43%) | 35/59<br>(59%) |
| Do you smoke cigarettes? | Yes | 20/32<br>(63%) | 17/27<br>(63%) | 27/41<br>(66%) | 27/48<br>(56%) | 28/46<br>(61%) | 23/39<br>(59%) | 36/55<br>(65%) | 35/54<br>(65%) | 37/54<br>(69%) | 36/59<br>(61%) |

**Table 2.** Proportion of children with hearing loss by category (normal, mild, moderate, severe), hearing loss above 23.3 dB, and mean hearing loss (dB, SD) at ages 12 (baseline), 18, 24, 30, and 36 months, by booster dose (difference, 95%CI, p value). (N=461)

| Visit age (months) | Hearing response category or response level (4fa dB, SD) | Total | +P | +S | DIFF +P - +S | 95%CI | P value |
| --- | --- | --- | --- | --- | --- | --- | --- |
| **12** | tested | 60/261(23%) | 32/131 (24%) | 28/130 (22%) | 3% | -7, 13 | 0.66 |
|  | none | 15 (25%) | 7/32 (22%) | 8/28 (29%) | -7% | -29, 15 | 0.57 |
|  | mild | 20 (33%) | 11 (34%) | 9 (32%) | 2% | -22, 26 | 1.00 |
|  | moderate | 25 (42%) | 14 (44%) | 11 (39%) | 4% | -20, 29 | 0.80 |
|  | >23.3 (3fa) | 40 (67%) | 22 (69%) | 18 (64%) | 4% | -19, 28 | 0.79 |
|  | Mean dB (SD) | 29.5 (8.4) | 29.5 (8.2) | 29.5 (8.8) | 0.02 dB | -4.4, 4.4 | 1.00 |
| **18** | tested | 91/257(35%) | 42/129 (33%) | 49/128 (38%) | -6% | -17, 6 | 0.36 |
|  | none | 23/91 (25%) | 15 (36%) | 8 (16%) | 19% | 2, 37 | 0.05 |
|  | mild | 39 (43%) | 18 (43%) | 21 (43%) | 0% | -20, 20 | 1.00 |
|  | moderate | 29 (32%) | 9 (21%) | 20 (41%) | -19% | -38, -1 | 0.07 |
|  | >23.3 (3fa) | 63 (69%) | 26 (62%) | 37 (76%) | -14% | -33, 5 | 0.18 |
|  | Mean dB (SD) | 28.0 (7.9) | 26.2 (6.9) | 29.6 (8.4) | **-3.4 dB** | -6.7, -0.2 | **0.04** |
| **24** | tested | 86/98 (88%) | 47/48 (98%) | 39/50 (78%) | 20% | 8, 32 | 0.004 |
|  | none | 19 (22%) | 11 (23%) | 8 (21%) | 3% | -15, 20 | 0.80 |
|  | mild | 38 (44%) | 21 (45%) | 17 (44%) | -1% | -20, 22 | 1.00 |
|  | moderate | 29 (34%) | 15 (32%) | 14 (36%) | -3% | -23, 18 | 1.00 |
|  | >23.3 (3fa) | 61 (71%) | 31 (66%) | 30 (77%) | -11% | -30, 8 | 0.34 |
|  | Mean dB (SD) | 28.9 (9.4) | 27.7 (8.1) | 30.3 (10.7) | -2.6 dB | -6.7, 1.4 | 0.2 |
| **30** | tested | 111/111 (100%) | 55/55 (100%) | 56/56 (100%) | 0% | na | na |
|  | none | 35 (34%) | 24 (44%) | 18 (32%) | 12% | -6, 29 | 0.24 |
|  | mild | 32 (29%) | 16 (29%) | 16 (29%) | 0% | -16, 17 | 1.00 |
|  | moderate | 37 (33%) | 15 (27%) | 22 (39%) | -12% | -30, 6 | 0.29 |
|  | >23.3 (3fa) | 56 (50%) | 26 (47%) | 30 (54%) | -6% | -25, 12 | 0.57 |
|  | Mean dB (SD) | 26.9 (9.4) | 26.0 (8.7) | 27.8 (10.05) | -1.9 dB | -5.4, 1.7 | 0.30 |
| **36** | tested | 113/196 (58%) | 54/91 (59%) | 59/105 (56%) | 3% | -11, 17 | 0.67 |
|  | none | 52 (42%) | 26 (48%) | 25 (42%) | 6% | -14, 14 | 0.57 |
|  | mild | 39 (34%) | 20(37%) | 19 (32%) | 5% | -13, 22 | 0.69 |
|  | moderate | 22 (19%) | 8 (15%) | 14 (24%) | -9% | -23, 5 | 0.25 |
|  | severe | 1 (0.6%) | 0 (0%) | 1 (1.7%) | -1% | -3, 1 | 1.00 |
|  | >23.3 (3fa) | 49 (43%) | 22 (41%) | 27 (45%) | -4% | -22, 14 | 0.71 |
|  | Mean dB (SD) | 24.5 (10.4) | 22.8 (7.2) | 25.9 (12.5) | -3.1 dB | -7.0, 0.7 | 0.11 |

**Bold** indicates significant vaccine group difference p<0.05
+P: booster dose of PCV13 13-valent Pneumococcal conjugate vaccine
+S: booster dose of PHiD-CV10 10-valent Pneumococcal *H. influenzae* protein D conjugate vaccine
dB: decibel
SD: standard deviation
Hearing loss categories: none ≤ 20dB. Mild 21 to 30 dB. Moderate 31 to 60 dB. Severe 61 to 90 dB. Profound > 90 dB
4fa: average of up to 4 frequencies (500, 1000, 2000, and 4000 Hertz)
>23.3 (3fa): Hearing Australia threshold for hearing assistance
CI: Confidence Interval

There were no significant vaccine inter-group differences in characteristics of children in the hearing cohort at any age, other than at age 24 months when the +S group had a higher baseline proportion of siblings with “runny ears” (32%) than the +P group (13%), and at age 36 months when the +S group had a higher baseline proportion of breast feeding in infancy (73%) than the +P group (52%). (Table 1)

At baseline age 12 months, 32/131 (24%) and 28/130 (22%) children in the +P and +S groups respectively had hearing tests; hearing loss was similar in both groups (Table 2, Fig 2). At age 18 months, six months post booster dose, 42 and 49 children had hearing tests in the +P and +S groups, respectively; 36% and 16%, respectively had no hearing loss (difference 19% [95%CI 2, 37], p=0·05); 43% in each group had mild hearing loss, and 21% and 41%, respectively had moderate hearing loss (difference −19% [95%CI −38, −1], p=0·07) (Table 2, Fig 2). Mean hearing level was 26·2 dB in the +P group and 29·6 dB in the +S group (difference −3·4 dB ([95%CI −6·7, −0·2] p=0·04) (Table 2); 62% children in the +P group and 76% in the +S group met the Hearing Australia threshold for hearing assistance (mean hearing level (3fa) greater than 23·3 dB)[11] (difference −14%, [95%CI −33, 5] p=0·18).(Table 2)

**Fig 2:**
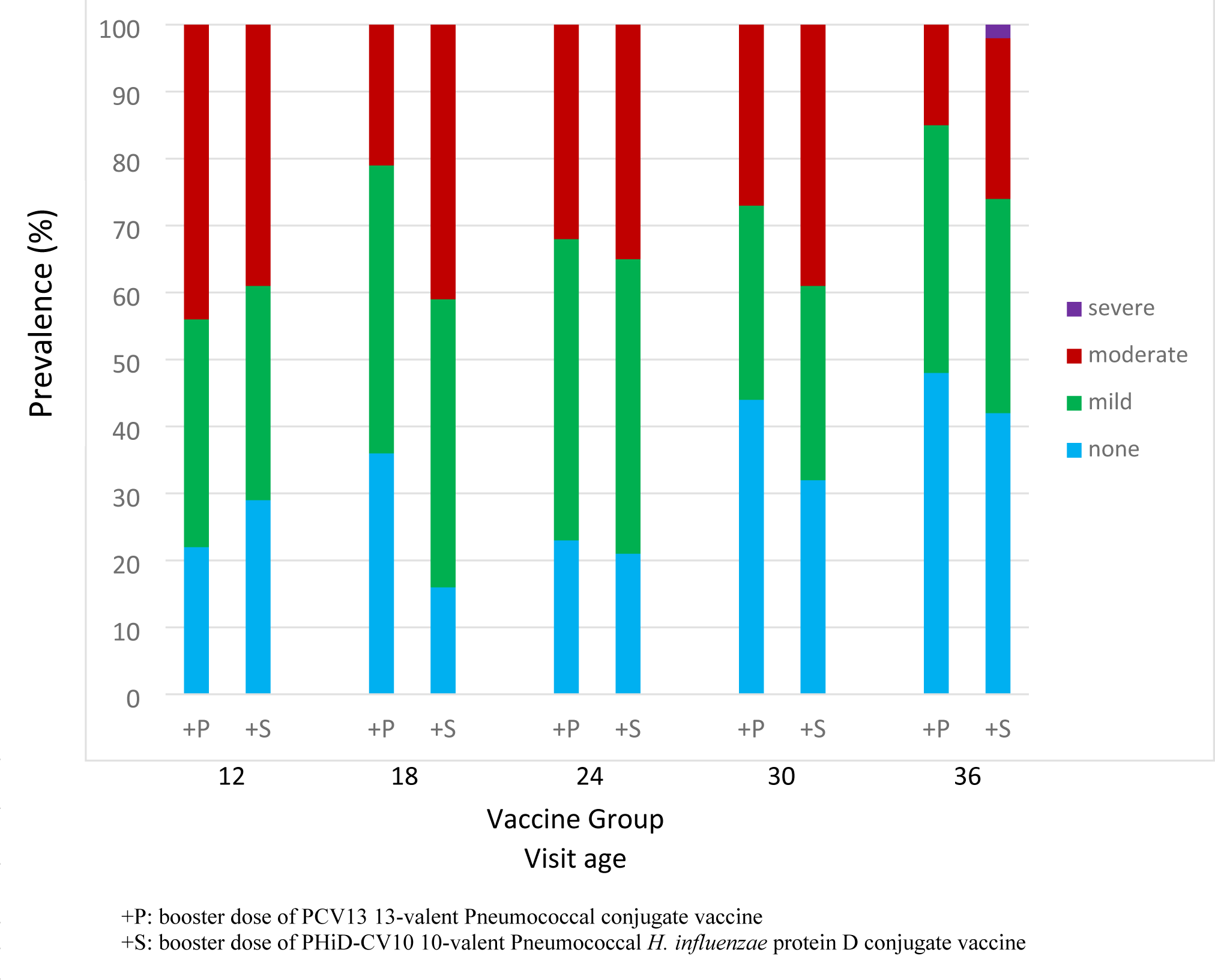
Hearing loss prevalence (normal, mild, moderate, severe) by booster dose, at ages 12 (baseline), 18 (primary endpoint), 24, 30, and 36 months.

At ages 24, 30, and 36 months, the proportion of children with no hearing loss was higher, mean hearing level and moderate hearing loss tended to be lower in the +P group. (Table 2, Fig 2)

Analysis stratified by allocation to three primary course schedules in PREVIX_COMBO indicated that the lower prevalence of moderate hearing loss in the +P group at age 18 months was present in all subgroups (differences −8% between _PPP+P and _PPP+S, −5% between _SSS+P and _SSS+S, and - 15% between SSSP+P and SSSP+S). The greatest difference was between SSSP+P (23%) and SSSP+S (38%) (difference −15% [95%CI −30, −0.1], p=0·07). (Table 3)

**Table 3.** Prevalence of hearing loss in the +P versus +S groups, stratified by allocation to three primary course PCV schedules (_PPP, _SSS, and SSSP) at age 18 months.

| Primary course | PPP |  |  | SSS |  |  | SSSP |  |  | TOTAL |
| --- | --- | --- | --- | --- | --- | --- | --- | --- | --- | --- |
| Booster dose | +P | +S |  | +P | +S |  | +P | +S |  |  |
| N | 80 | 63 |  | 85 | 91 |  | 65 | 76 |  | 460 |
| Category of hearing loss | n (%) | n (%) | Diff [95%CI]<br>p value | n (%) | n (%) | Diff [95%CI]<br>p value | n (%) | n (%) | Diff [95%CI]<br>p value |  |
| None (< 20dB) | 26 (33%) | 17 (27%) | 6% [-10, 21]<br>0.58 | 31 (36%) | 25 (27%) | 9% [-5, 23]<br>0.26 | 26 (40%) | 25 (33%) | 7% [-9, 23]<br>0.48 | 150 (33%) |
| Mild (21-30 dB) | 31 (39%) | 23 (37%) | 2% [-14, 18]<br>0.86 | 31 (36%) | 37 (41%) | -4% [-19, 10]<br>0.64 | 24 (37%) | 22 (29%) | 8% [-7, 24]<br>0.37 | 168 (37%) |
| Moderate (31-60 dB) | 23 (29%) | 23 (37%) | -8%<br>[-23, 8]<br>0.37 | 23 (27%) | 29 (32%) | -5%<br>[-18, 9]<br>0.51 | 15 (23%) | 29 (38%) | -15% [-30, -0.1]<br>0.069 | 142 (31%) |

Fluctuating hearing loss was examined using data from children who had hearing tests at two or more consecutive 6-monthly assessments. Overall, at initial assessment the proportion of hearing tests that found no hearing loss, mild, or moderate hearing loss was similar (35%, 36%, 29%, respectively) to 6-monthly follow-up (38%, 36%, 25%, respectively). However, transitions in hearing status were more dynamic than revealed by point prevalence: of 73 children with no hearing loss initially, 51% had no hearing loss at follow-up, 37% transitioned to mild-, and 12% to moderate-hearing loss. Of 76 children with mild hearing loss initially, 32% transitioned to no hearing loss, 42% had persisting mild hearing loss, 26% transitioned to moderate hearing loss, and 1% to severe hearing loss. Of 61 children with moderate hearing loss initially, 33% transitioned to no hearing loss, 28% transitioned to mild hearing loss, and 39% had persisting moderate hearing loss. (Table 4)

**Table 4.** Transitions between consecutive 6-monthly hearing assessments.

| Hearing loss category after 6 months: | Normal: <20 dB | Mild: 21 to 30 dB | Moderate: 31 to 60 dB | Severe: 61 to 90 dB | Total |
| --- | --- | --- | --- | --- | --- |
| Hearing loss category at initial assessment: |  |  |  |  |  |
| Normal: <20 dB | 37/73<br>51% | 27<br>37% | 9<br>12% | 0 | 73 (35%) |
| Mild: 21 to 30 dB | 24/77<br>31% | 32<br>42% | 20<br>26% | 1<br>(1%) | 77 (36%) |
| Moderate: 31 to 60 dB | 20<br>33% | 17<br>28% | 24<br>39% | 0 | 61 (29%) |
| Total | 81<br>(38%) | 76<br>(36%) | 53<br>(25%) | 1<br>(0.5%) | 211<br>(100%) |

## Discussion

To our knowledge this is the first study globally to evaluate hearing loss outcomes in an RCT of head-to-head or mixed schedule PCVs. This is also the first prospective longitudinal community-based study of scheduled hearing tests in First Nations children from age 12 to 36 months. A literature search for studies of either PCV13 or PHiD-CV10 that report either otitis media or hearing-related outcomes,[13] was repeated for the period 2019 to 2023; of 54 identified studies, none reported hearing outcomes. A recent 2020 Cochrane review of PCVs for prevention of acute otitis media did not report hearing outcomes.[15] Other head-to-head RCTs compare immunogenicity of different PCV schedules or formulations[16, 17] and impact on pneumococcal[18] and NTHi carriage[16] but impacts on hearing have not been included.

Unexpectedly, this report focussing on hearing outcomes found that, contrary to our hypotheses, the children assessed in the +P group had better hearing compared to the +S group at age 18 months. The −3 dB difference in mean hearing level is of unknown clinical value. However, the −19% difference in prevalence of moderate hearing loss and gain of 19% having no hearing loss is likely to benefit many First Nations children, particularly if this is sustained to age 36 months. In addition, around two-thirds of the children assessed at age 18 months met Hearing Australia criteria for hearing assistance,[11] and the proportion was again lower in the +P group at age 18 months (62% versus 76%). At later ages vaccine group differences in hearing persisted. Analysis stratified by the primary course head-to-head and combination schedules (the PREVIX_COMBO RCT)[5] suggested that slightly better hearing in the +P group was present across all primary schedules and was greatest in the SSSP+P versus SSSP+S comparison. These unique findings are difficult to explain and should be interpreted with caution as not all randomised participants received hearing assessments.

A limitation of our study is small sample size for this secondary outcome,[19] although there were no boost-group differences in rate of loss-to-follow-up, nor participant characteristics or risk factors at any age. The high event rate somewhat compensates for loss-to-follow-up and may increase confidence in the apparent vaccine group difference in hearing detected at primary endpoint age 18 months. Given the paucity of published data from vaccine trials to support or refute our findings, further investigation of biological plausibility is warranted, particularly as higher valency vaccines become available.[20]

Our systematic review identified an almost complete absence of published contemporary (post PCVera) data on the prevalence and persistence of hearing loss in very young First Nations children.[21] In addition, data from the NT hearing outreach services show that only 11% of services (either audiology, Clinical Nurse Specialist, or ear, nose and throat (ENT) teleotology) were provided to children under age 3 years.[22] Of 1,741 First Nations children who received audiology services, 56 (3%) were under age 3 years and of these 17 (31%) had some level of hearing loss.

The systematic review[21] and national otitis media guidelines[13] also highlight the paucity of prevalence studies and absence of published intervention studies that address this crisis. In 2022 and 2023, the first community-based prevalence studies of hearing loss in First Nations children living in urban settings were published; one identified that at age 12 months, 69% of 67 children tested had hearing loss, including 24 (36%) with moderate hearing loss.[23] The second, larger study which excluded children under age 3 years obtained hearing data from 1087 (median age 8·2 years) of 1669 children enrolled; 279 (25·7%) had hearing loss.[24] Neither study was an evaluation of hearing loss rehabilitation or prevention strategies.

Using combined vaccine group data, our key findings identified for the first time the high prevalence of persistent and fluctuating mild-moderate hearing loss throughout early childhood, from 75% at age 12 months to 53% at age 36 months. In particular the high prevalence of moderate hearing loss, defined by the World Health Organization as disabling for children,[10] which was 42% at age 12 months, declining slowly to 19% at age 36 months.

These findings place many children in need of hearing assistance according to the Hearing Australia threshold of ≥ 23·3 dB.[11] Also, according to national OM Guidelines,[13] at ages 12 to 30 months, around 30% children met criteria for ENT consultation, based on the mean hearing level in the better ear of >30 dB.

Transitions in level of hearing loss between consecutive 6-monthly visits, despite stable prevalence rates at each timepoint, confirm the fluctuating nature of conductive hearing loss in this age group. This has practical implications that support the shift from screening programs to surveillance, commencing within first year of life.[25]

A strength of our study is that all hearing tests were performed in sound-treated facilities by paediatric audiologists at scheduled study visits, regardless of clinical presentation. However, the majority of hearing tests were VROA for which the minimum test level varies according to test environment and child. If the minimum test level available was 20 to 25 dB, children with a true hearing level below test limits could be misidentified as having mild hearing loss using our study definitions, producing a false positive and over-inflating the prevalence of mild hearing loss as determined by VROA.

From primary health care to specialist services there are huge shortfalls that disrupt a child’s hearing care pathway.[26] At December 2022, there were 3,265 Indigenous children and young people on the wait list for fly-in/fly-out audiology outreach services and 2,284 on the ENT teleotology wait list.[22] As mentioned, in 2022 very few outreach services were for children under age 3 years, and ENT surgeries were rarely performed (4% of all ENT services were for grommets).[22]

This report of the extraordinarily high prevalence of preventable hearing loss in First Nations children highlights the need for effective social and public health reforms with greater investment in interventions that target social determinants, namely more preventative models.[27] Others have demonstrated the inequalities in ENT service provision between Aboriginal and non-Aboriginal children, attributed to socioeconomic status and geographical isolation.[28] In the US, social deprivation limits children’s access to appropriate medical and surgical treatment for their ear disease and increases odds of severe complications.[29] Unfortunately studies of interventions that address social determinants will be difficult and few that report otitis media outcomes have been published or show a benefit.[30] Our research program is currently evaluating a primary health care workforce enhancement strategy (the Hearing for Learning Initiative) of remote on-country training and employment of ear health facilitators which could provide a sustainable, culturally appropriate and safe model to detect and prevent disabling hearing loss, and support children who have hearing problems.[31, 32]

## Data Availability

Availability of data/Data sharing Data collected for the study that underlie the results reported in this Article, including individual participant data and a data dictionary defining each field in the set, will be made available after deidentification. No additional, related documents will be available, and the study protocols have been published. Data will be available up to 3 years after publication of this Article, upon request to the corresponding author. Data will be shared with investigators whose proposed use of the data has been approved by the ethics committees of NT Health, Menzies School of Health Research, WA Department of Health, and WA Aboriginal Health Ethics Committee, and approved by the Menzies’ Child Health Division’s Australian First Nations Reference Group, for analyses that meet criteria for excellence in research with Aboriginal and Torres Strait Islander people, and with an institutional-signed and investigator signed data sharing research agreement.

## DECLARATIONS

### Ethics approval and consent to participate

Ethical approval has been obtained from Human Research Ethics Committees of the Northern Territory Department of Health and Menzies School of Health Research (NHMRC Reg no: EC00153), the Central Australian HREC (NHMRC Reg no: EC00155) and West Australian Aboriginal Health Ethics Committee (WAAHEC-377-12/2011). Parents or guardians provided signed informed consent for their infant’s participation.

### Availability of data/Data sharing

Data collected for the study that underlie the results reported in this Article, including individual participant data and a data dictionary defining each field in the set, will be made available after de-identification. No additional, related documents will be available, and the study protocols have been published. Data will be available upon request to the Senior Data Manager at the Menzies School of Health Research. Data will be shared with investigators whose proposed use of the data has been approved by the ethics committees of NT Health, Menzies School of Health Research, WA Department of Health, and WA Aboriginal Health Ethics Committee, and approved by the Menzies’ Child Health Division’s Australian First Nations Reference Group, for analyses that meet criteria for excellence in research with Aboriginal and Torres Strait Islander people, and with an institutional-signed and investigator signed data sharing research agreement.

### Competing interests (use author initials)/Declaration of interests

**AJL** received funds from NHMRC paid to the institution, and GSK provided materials for immunogenicity assays. AJL received funds from Merck Sharp and Dohme for analysis of pneumococcal carriage, payment to institution. **ABC** served as advisor on a Data Safety Monitoring Board for an unlicensed vaccine (GlaxoSmithKline) and an unlicensed monoclonal antibody (AstraZeneca), was an adviser on an unlicensed molecule for chronic cough (Merck); and has multiple project grants and a Centre of Research Excellence relating to various aspects of bronchiectasis in children from the National Health and Medical Research Council. ABC received Royalties or licences as an author of cough and bronchiectasis topics, and Partial reimbursement for airfares as a speaker for European Respiratory Society. All payments were to the institution. **PM** served on a data safety and monitoring board for the Novavax COVID-19 vaccine. **JB** provided a report to MSD Australia. All other authors (DL, CB-J, HS-V, MB, MC, PL, PSM, PT,) declare no competing interests.

### Data safety monitoring

The study was overseen by an independent Data Safety and Monitoring Board (iDSMB).

### Author contributions

AJL (Principal Investigator, PI) conceived the study, led funding applications, obtained HREC approval and other regulatory approvals, undertook consultations, reporting and had overseen day to day management and implementation of the trials, managed, had direct access to and verified the reported data, analysed and interpreted the data, created tables and figures, wrote the first draft and final version of manuscript. NW managed the trial, participant recruitment and retention, specimen collection, reported to Ethics committees and data safety monitoring board, managed quality of data and read the final version of the manuscript. BA assisted participant recruitment and retention, specimen collection, managed quality of data and read the final version of the manuscript. JB managed microbiology and serology collections, data base and data quality, and read the final version of the manuscript. EKM advised on study design, assisted with funding application, participated in investigator meetings, advised on risk management and read the final version of the manuscript. MS advised on study design, assisted with funding application, participated in investigator meetings, advised on risk management and read the final version of the manuscript. PJT advised on study design, assisted with funding application, participated in investigator meetings, advised on risk management and read the final version of the manuscript. PM advised on study design, assisted with funding application, participated in investigator meetings, advised on risk management and read the final version of the manuscript. HS-V advised on study design, assisted with funding application, participated in investigator meetings, advised on microbiology protocols, and reviewed the final version of the manuscript. SS advised on study design, assisted with funding application, participated in investigator meetings, advised on laboratory protocols, particularly microbiology, and reviewed the final version of the manuscript. VO participated in investigator meetings, wrote the final statistical analysis plans, had direct access to and verified the reported data, generated tables and figures and read the final version of the manuscript. MC advised on study design, assisted with funding application, participated in investigator meetings, wrote the initial statistical analysis plans in the protocols, analysed data, generated tables and figures and read the final version of the manuscript. DL advised on study design, assisted with funding application, participated in investigator meetings, advised on tables and figures and read the final version of the manuscript. CB-J advised on interpretation of audiology data and read the final version of the manuscript. MB advised on study design, assisted with funding application, participated in investigator meetings, advised on tables and figures and read the final version of the manuscript. PL advised on laboratory protocols, particularly immunogenicity, and read the final version of the manuscript. RMA advised on study design, assisted with funding application, participated in investigator meetings, advised on tables and figures and read the final version of the manuscript. TS advised on study design, assisted with funding application, participated in investigator meetings, advised on tables and figures and read the final version of the manuscript. VK advised on study design, assisted with funding application, participated in investigator meetings, advised on tables and figures and read the final version of the manuscript. JC advised on study design, assisted with funding application, participated in investigator meetings, advised on tables and figures and read the final version of the manuscript. ABC advised on study design, assisted with funding application, participated in investigator meetings, advised on tables and figures and read the final version of the manuscript. PSM advised on study design, assisted with funding application, participated in investigator meetings, advised on risk management and provided day to day supervision of clinical training, and read the final version of the manuscript.

## Acknowledgements

Sources of funding were the Australian NHMRC (National Health and Medical Research Council (GNT605810, GNT1046999, GNT1120353) and GlaxoSmithKline (donation of reagents). AJL was not paid by any agency to write this article. We thank the many families living in remote communities in northern Australia who participated in the PREVIX_COMBO and PREVIX_BOOST randomised controlled trials. We thank the many research nurses, audiologists, laboratory staff, and others who worked on the trials.

